# Higher mortality associated with the SARS-CoV-2 Delta variant in the Western Cape, South Africa, using RdRp target delay as a proxy

**DOI:** 10.1101/2021.10.23.21265412

**Authors:** Hannah Hussey, Mary-Ann Davies, Alexa Heekes, Carolyn Williamson, Ziyaad Valley-Omar, Diana Hardie, Stephen Korsman, Deelan Doolabh, Wolfgang Preiser, Tongai Maponga, Arash Iranzadeh, Susan Engelbrecht, Sean Wasserman, Neshaad Schrueder, Linda Boloko, Greg Symons, Peter Raubenheimer, Abraham Viljoen, Arifa Parker, Cheryl Cohen, Waasila Jassat, Richard Lessells, Robert J Wilkinson, Andrew Boulle, Nei-yuan Hsiao

## Abstract

A novel proxy for the Delta variant, RNA-dependent RNA polymerase target delay in the Seegene Allplex™ 2019-nCoV PCR assay, was associated with higher mortality (adjusted Odds Ratio 1.45 [95%CI 1.13-1.86]), compared to presumptive Beta infection, in the Western Cape, South Africa (April-July 2021). Prior diagnosed infection and vaccination were protective.

## Introduction

To date, the Western Cape Province of South Africa, has experienced three large waves of SARS-CoV-2 infections. The first wave caused by early ancestral clades of SARS-CoV-2 peaked in June 2020, the second wave with the Beta variant (B.1.351) peaked in December 2020 and the third most recent wave caused by the Delta variant (B.1.617.2) peaked in August 2021 (1).

Delta has become the dominant variant globally, and its increased clinical severity compared to other variants is suspected, although not fully documented or understood (2–6). Relatively high vaccination coverage, however, complicates the interpretation of these findings. Delta has also mostly been compared to the Alpha variant, and not within a sub-Saharan African setting.

The start of the third wave of COVID-19 in the Western Cape Province was characterized by a rapid transition from the previously dominant Beta variant to Delta (1). We used a novel proxy marker for Delta, namely RNA-dependent RNA polymerase (RdRp) target delay (RTD) in PCR positive samples, to assess mortality associated with Delta, compared to Beta, in our population which had relatively low levels of vaccine coverage as well as a high prevalence of comorbidities, including HIV and tuberculosis.

## Methods

### RdRP target delay

RTD, defined as a difference in cycle threshold value of >3.5 in the RdRp relative to E gene target in the Seegene Allplex™ 2019-nCoV PCR assay, has recently been described (7). This phenomenon is due to the G15451A mutation found in Delta, resulting in a RdRp primer mismatch. When evaluated against genomic data, this method had a sensitivity of 93.6% and specificity of 89.7% in detecting Delta (7).

### Population and Statistical analysis

We included all COVID-19 cases diagnosed on the Seegene Allplex ™ 2019-nCoV diagnostic PCR assay in the Western Cape public sector from 1 April to 31 July 2021, a period when both Beta and Delta were co-circulating (Appendix Figure 1). Alpha and non-variant of concern lineages accounted for a negligible number of infections at this time (1). Follow-up ended on 31 August 2021. Approximately 70% of the Western Cape population uses the public sector for health services and the Western Cape Provincial Health Data Centre (PHDC) collates all available electronic health data on these patients. Use of this data in COVID-19 analyses has been previously described (8). COVID-19 deaths were defined as deaths within 28 days of diagnosis of COVID-19 or 14 days after discharge from hospital (where hospital admission occurred within 21 days of the COVID-19 diagnosis), without a non-COVID-19 cause of death recorded. Out-of-facility deaths were determined by civil identifier linkage of patients with COVID-19 diagnoses to the national population register.

We undertook logistic regression to determine the association between RTD and mortality, adjusted for age, sex, known comorbidities, prior diagnosed infection, vaccination status at time of diagnosis, sub-district of residence, month of diagnosis and hospital admission pressure (number of public sector admissions in the calendar week of diagnosis). Prior diagnosed infections were defined as a positive COVID-19 test more than 90 days prior to the current test and classified into those with their first infection in the first wave (1 March – 30 September 2020) or second wave (1 October 2020 – 31 March 2021). Vaccination status at time of COVID-19 diagnosis was determined by linking COVID-19 cases with the national Electronic Vaccination Data System through national identifiers in the PHDC. Complete vaccination was defined ≥28 days post-vaccination with Janssen/Johnson & Johnson (Ad26.COV2.S), or ≥14 days post second dose of Pfizer–BioNTech (BNT162b2). Patients were deemed partially vaccinated from the day after their (first) vaccine dose until meeting criteria for complete vaccination.

The study was approved by the University of Cape Town Research Ethics Committee (HREC 460/2020).

## Results

We included 11 355 cases tested using the Seegene Allplex ™ assay, which were 22% of all positive test results in the province in that time period (Appendix Table 1). The median age was 43 years (interquartile range [IQR] 32-55) and 44% were male. RTD was present in 9106 (80%) (Table 1). Patient characteristics were similar in those with and without RTD. There was, however, a difference in cycle threshold (Ct) values (average of the E and N gene targets); those with RTD had lower median Ct values (26.1, IQR 22.2-30.9, vs 32.7 IQR 25.6-37.6).

**Table 1:**
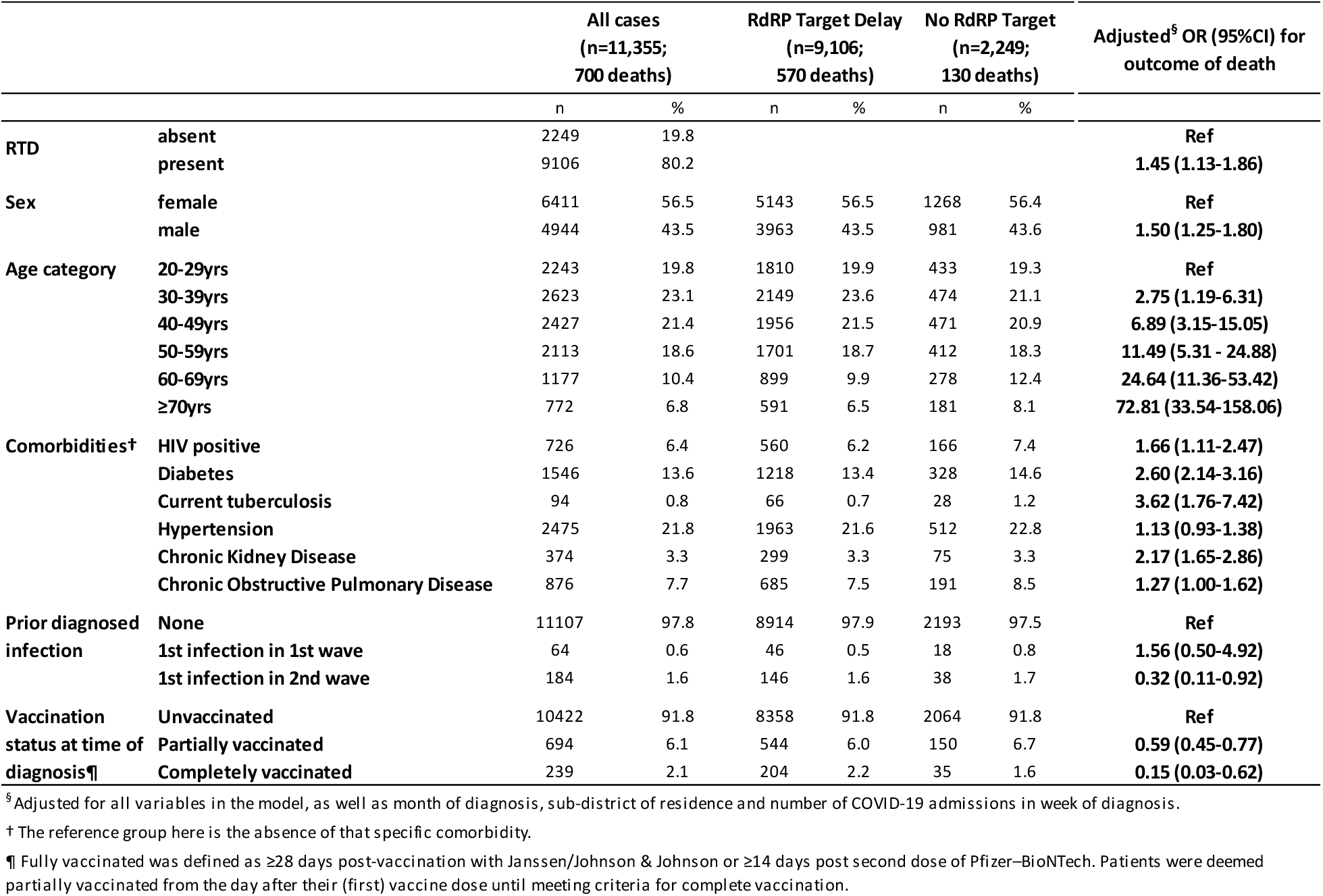
Characteristics of all COVID-19 cases (Seegene Allplex™ 2019-nCoV assay positive) in the Western Cape, April - July 2021, by presence of RdRp target delay (RTD), and results from logistic regression of associations with the outcome of death.

|  |  | All cases<br>(n=11,355;<br>700 deaths) |  | RdRP Target Delay<br>(n=9,106;<br>570 deaths) |  | No RdRP Target<br>(n=2,249;<br>130 deaths) |  | Adjusted<sup>§</sup> OR (95%CI) for<br>outcome of death |
| --- | --- | --- | --- | --- | --- | --- | --- | --- |
|  |  | n | % | n | % | n | % |  |
| RTD | absent | 2249 | 19.8 |  |  |  |  | Ref |
|  | present | 9106 | 80.2 |  |  |  |  | 1.45 (1.13-1.86) |
| Sex | female | 6411 | 56.5 | 5143 | 56.5 | 1268 | 56.4 | Ref |
|  | male | 4944 | 43.5 | 3963 | 43.5 | 981 | 43.6 | 1.50 (1.25-1.80) |
| Age category | 20-29yrs | 2243 | 19.8 | 1810 | 19.9 | 433 | 19.3 | Ref |
|  | 30-39yrs | 2623 | 23.1 | 2149 | 23.6 | 474 | 21.1 | 2.75 (1.19-6.31) |
|  | 40-49yrs | 2427 | 21.4 | 1956 | 21.5 | 471 | 20.9 | 6.89 (3.15-15.05) |
|  | 50-59yrs | 2113 | 18.6 | 1701 | 18.7 | 412 | 18.3 | 11.49 (5.31 - 24.88) |
|  | 60-69yrs | 1177 | 10.4 | 899 | 9.9 | 278 | 12.4 | 24.64 (11.36-53.42) |
|  | ≥70yrs | 772 | 6.8 | 591 | 6.5 | 181 | 8.1 | 72.81 (33.54-158.06) |
| Comorbidities† | HIV positive | 726 | 6.4 | 560 | 6.2 | 166 | 7.4 | 1.66 (1.11-2.47) |
|  | Diabetes | 1546 | 13.6 | 1218 | 13.4 | 328 | 14.6 | 2.60 (2.14-3.16) |
|  | Current tuberculosis | 94 | 0.8 | 66 | 0.7 | 28 | 1.2 | 3.62 (1.76-7.42) |
|  | Hypertension | 2475 | 21.8 | 1963 | 21.6 | 512 | 22.8 | 1.13 (0.93-1.38) |
|  | Chronic Kidney Disease | 374 | 3.3 | 299 | 3.3 | 75 | 3.3 | 2.17 (1.65-2.86) |
|  | Chronic Obstructive Pulmonary Disease | 876 | 7.7 | 685 | 7.5 | 191 | 8.5 | 1.27 (1.00-1.62) |
| Prior diagnosed<br>infection | None | 11107 | 97.8 | 8914 | 97.9 | 2193 | 97.5 | Ref |
|  | 1st infection in 1st wave | 64 | 0.6 | 46 | 0.5 | 18 | 0.8 | 1.56 (0.50-4.92) |
|  | 1st infection in 2nd wave | 184 | 1.6 | 146 | 1.6 | 38 | 1.7 | 0.32 (0.11-0.92) |
| Vaccination<br>status at time of<br>diagnosis¶ | Unvaccinated | 10422 | 91.8 | 8358 | 91.8 | 2064 | 91.8 | Ref |
|  | Partially vaccinated | 694 | 6.1 | 544 | 6.0 | 150 | 6.7 | 0.59 (0.45-0.77) |
|  | Completely vaccinated | 239 | 2.1 | 204 | 2.2 | 35 | 1.6 | 0.15 (0.03-0.62) |

<sup>§</sup> Adjusted for all variables in the model, as well as month of diagnosis, sub-district of residence and number of COVID-19 admissions in week of diagnosis.
† The reference group here is the absence of that specific comorbidity.
¶ Fully vaccinated was defined as ≥28 days post-vaccination with Janssen/Johnson & Johnson or ≥14 days post second dose of Pfizer–BioNTech. Patients were deemed partially vaccinated from the day after their (first) vaccine dose until meeting criteria for complete vaccination.

There were 700 deaths altogether, with a case fatality rate of 6.3% in those with RTD, compared to 5.8% in those without. After adjusting for all covariates RTD was significantly associated with death among cases (adjusted odds ratio [aOR] 1.45 [95% confidence interval [CI] 1.13-1.86]) and among those admitted to hospital (aOR 1.39 [95% CI 1.03-1.88]) (Appendix Table 2).

Prior diagnosed infection with a first infection in the Beta dominated second wave (vs no prior diagnosed infection), was protective against death (aOR 0.32 [95% CI 0.11-0.92]), whereas prior diagnosed infection with a first infection in the first wave was not (aOR: 1.56 [95% CI 0.50-4.92]). Vaccination was protective against death. The aORs (95% CI) for partial and full vaccination were 0.59 (0.45-0.77) and 0.15 (0.03-0.62) respectively.

In a stratified analysis the association between RTD and death was similar in both those aged <50 and ≥50 years (aOR [95%CI] of 1.32 [0.77-2.26] and 1.44 [1.09-1.91] respectively) (Appendix Table 2).

## Discussion

RTD, a proxy marker for the Delta variant, was associated with an increased risk for death compared to presumptive Beta variant infection, while prior diagnosed infection (with first infection in the second wave) and vaccination were strongly protective.

The increased transmissibility of Delta is well established, and may be due to its higher viral load (6). In our analysis, specimens with RTD had lower Ct values than those without, suggesting a higher viral load. This, along with immune evasion by Delta, could contribute to the increased disease severity seen (9). Nonetheless, our results need to be interpreted in the context of our setting where not all COVID-19 cases would have accessed testing. Delta often results in mild symptoms, and during wave surges, as was the case from 15 June 2021 until the end of the analysis period, in the public sector only those aged ≥45 years or with comorbidities or requiring hospital admission would be tested. The interpretation of our results is therefore that among those who access a PCR test, the Delta variant is associated with worse outcomes. While it is unclear if this finding can be extrapolated to all those with COVID-19 in our setting, similar findings from countries with more widespread testing support the increased clinical severity of Delta.

Hospital admission was more likely with Delta compared to Alpha in England (aHR 2.26 [95%CI 1.32-3.89]) (2) and Denmark (RR 2.83 [95%CI 2.02-3.98]) (3). In Singapore, infection with Delta and Beta vs. wild-type SARS-CoV-2 were both associated with a composite outcome of oxygen use, intensive care admission and death (aOR 4.9 [95%CI 1.43-30.78] and 1.69 [95%CI 0.19-14.69] respectively) (4). Delta (vs wild-type) was associated with mortality in Canada (aOR 2.32 [95% CI 1.47-3.30]) as were N501Y-positive variants (Alpha, Beta, Gamma) (aOR 1.51 [95%CI 1.30-1.74]) (5). However, most of these countries had relatively high COVID-19 vaccination coverage rates by the time the Delta variant became dominant there (10). Since most severe COVID-19 cases would be in unvaccinated people, differences between the vaccinated and the unvaccinated population might confound associations with variant infection and disease severity. In contrast, in the Western Cape by 31 July 2021, only 5.0% of all adults, mostly in older age groups, were fully vaccinated (10). In addition, most of these studies compared the outcomes of Delta to Alpha. The Alpha variant has been shown to have worse clinical outcomes compared to wild-type, with an aHR for death of 1.73 (95%CI 1.41 - 2.13) (11).

This study has several limitations. First, we could only include cases tested on the Seegene Allplex™ 2019-nCoV assay, excluding cases diagnosed by other PCR methods or antigen testing. However, the included cases are mostly representative of all diagnosed PCR cases in the Western Cape (Appendix Table 1). Second, while RTD is a reliable proxy marker for Delta, it is not as accurate as whole genome sequencing, and misclassification may have diluted the effect of Delta. Third, we could only assess the effect of prior laboratory-confirmed COVID-19 infection and seroprevalence studies suggest considerably higher numbers were infected, even after the first wave, and that prior infection prevalence differed by sub-district of residence (12). While we did adjust for sub-district, the absence of a protective effect of prior infection in the first wave may be due to insufficient numbers of those infections being diagnosed. Finally, although we adjusted for COVID-19 hospital admissions to account for escalating service pressure during the wave surge, we could not adjust for non-COVID-19 admissions that might have added to pressure on facilities and contributed to mortality.

## Conclusion

RTD, a useful proxy for infection with Delta, is associated with an increased risk of mortality amongst those who were tested for COVID-19 in our setting.

## Data Availability

All data produced in the present study are available upon reasonable request to the authors

---

**Appendix: Figure 1.**
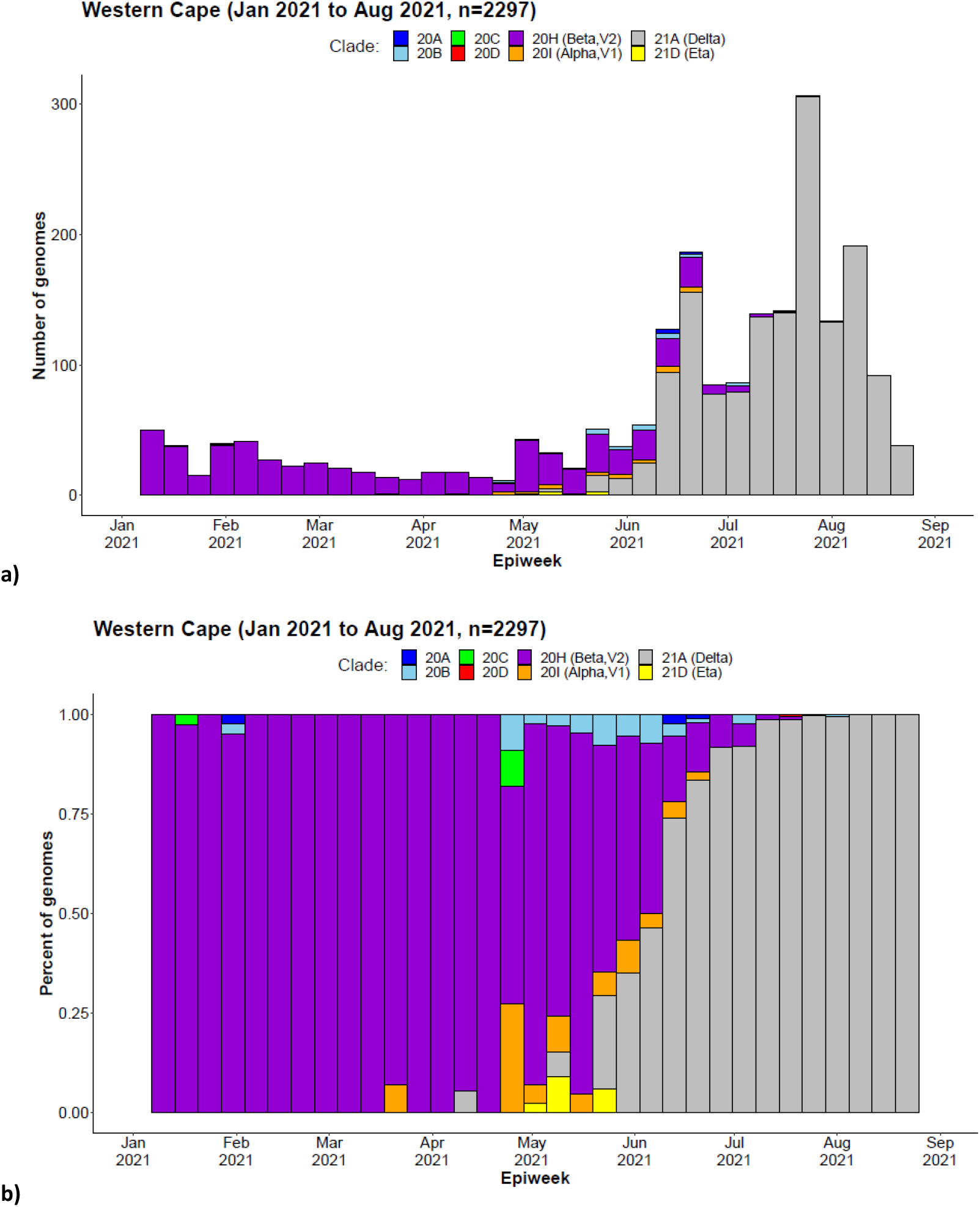
**Weekly frequency and distribution of SARS-CoV-2 variants circulating in the Western Cape Province, South Africa, 1 January to 31 August 2021: a) absolute count of genomes sequenced, b) proportion of genomes sequenced**. (7).

**Appendix: Table 1.**
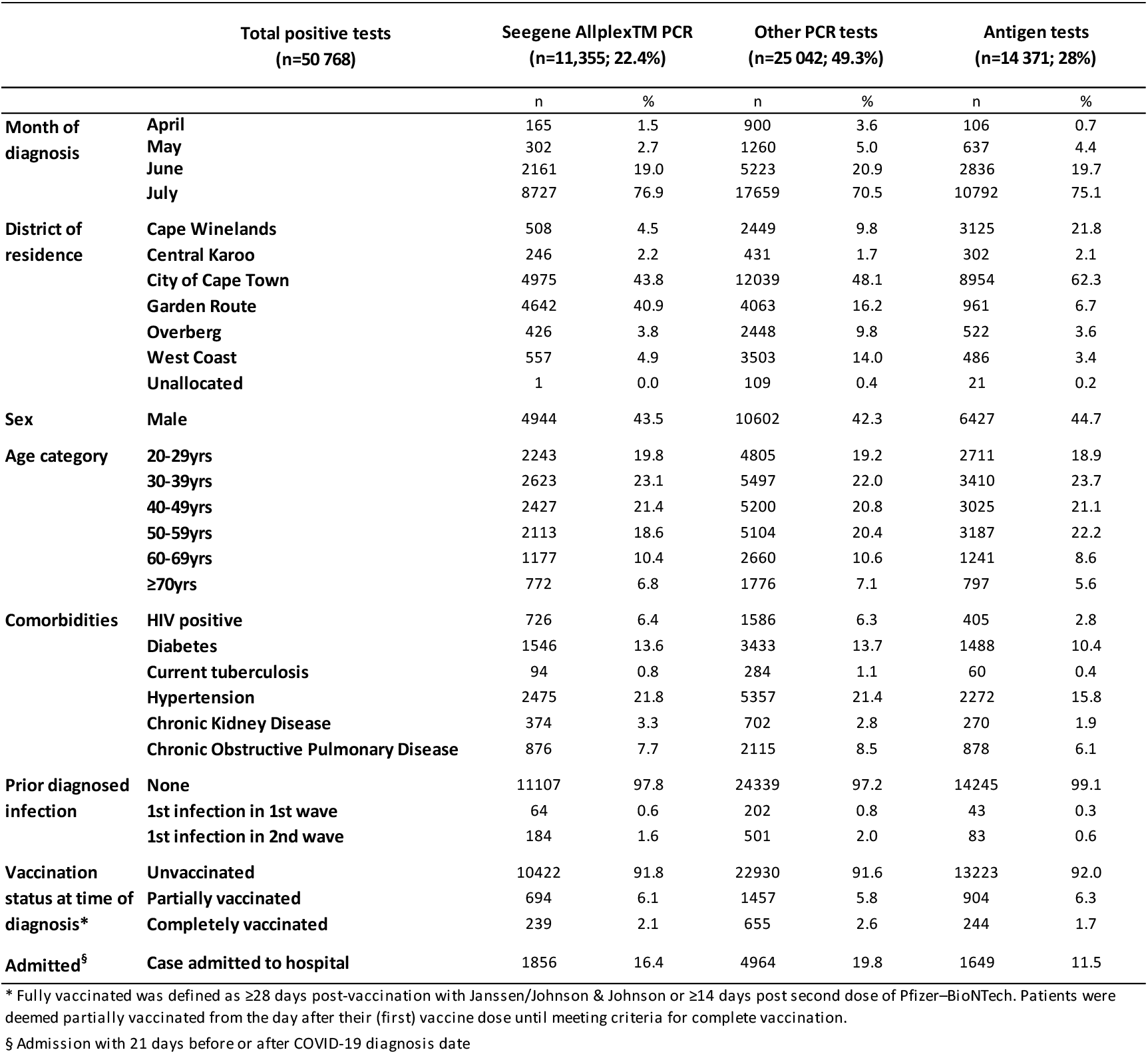
Patient characteristics by test type, Western Cape public sector, April to July 2021.

Patients tested on the Seegene Allplex™ PCR were similar in age, sex, known comorbidities, prior diagnosed infection and vaccination status to those who tested positive using other PCR assays. As different laboratories service different geographical regions there was a difference, however, in the district of residence. Patients who tested positive on antigen tests tended to be younger, have fewer comorbidities and fewer were admitted to hospital. This is in accordance to the Western Cape’s testing guidelines where PCR was preferred for hospital patients, while antigen testing was promoted at primary health care facilities.

**Appendix: Table 2.** Logistic regression for outcome of death in a) all positive Seegene Allplex™ cases, as per the original analysis; b) restricted only to those admitted to hospital; c) stratified by age into those aged under 50 and those aged 50 years.

|  |  | Adjusted<sup>§</sup> OR (95%CI) for outcome of death |  |  |  |
| --- | --- | --- | --- | --- | --- |
|  |  | All cases<br>(n=11 355; 700 deaths) | Only admitted cases*<br>(n=1 856; 612 deaths) | All cases < 50 years<br>(n=7 293; 113 deaths) | All cases ≥ 50 years<br>(n=4 062; 587 deaths) |
| RTD | absent | Ref | Ref | Ref | Ref |
|  | present | 1.45 (1.13-1.86) | 1.39 (1.03-1.88) | 1.32 (0.77-2.26) | 1.44 (1.09-1.91) |
| Sex | female | Ref | Ref | Ref | Ref |
|  | male | 1.50 (1.25-1.80) | 1.19 (0.95-1.47) | 1.16 (0.78-1.72) | 1.63 (1.33-2.00) |
| Age category | 20-29yrs | Ref | Ref | Ref |  |
|  | 30-39yrs | 2.75 (1.19-6.31) | 2.19 (0.81-5.93) | 2.49 (1.07-5.78) |  |
|  | 40-49yrs | 6.89 (3.15-15.05) | 5.91 (2.27-15.54) | 6.48 (2.92-14.37) |  |
|  | 50-59yrs | 11.49 (5.31 - 24.88) | 6.86 (2.66-17.67) |  | Ref |
|  | 60-69yrs | 24.64 (11.36-53.42) | 11.75 (4.56-30.28) |  | 2.16 (1.68-2.79) |
|  | ≥70yrs | 72.81 (33.54-158.06) | 25.93 (10.03-67.01) |  | 6.43 (4.96-8.33) |
| Comorbidities† | HIV positive | 1.66 (1.11-2.47) | 1.14 (0.69-1.88) | 2.21 (1.27-3.85) | 0.99 (0.53-1.85) |
|  | Diabetes | 2.60 (2.14-3.16) | 1.19 (0.95-1.49) | 2.58 (1.55-4.32) | 2.61 (2.11-3.22) |
|  | Current tuberculosis | 3.62 (1.76-7.42) | 1.57 (0.68-3.61) | 4.90 (1.90-12.65) | 2.38 (0.78-7.20) |
|  | Hypertension | 1.13 (0.93-1.38) | 0.95 (0.76-1.20) | 1.07 (0.63-1.82) | 1.16 (0.93-1.43) |
|  | Chronic Kidney Disease | 2.17 (1.65-2.86) | 1.76 (1.29-2.42) | 9.33 (4.20-20.72) | 1.87 (1.41-2.50) |
|  | Chronic Obstructive Pulmonary Disease | 1.27 (1.00-1.62) | 0.91 (0.69-1.19) | 1.95 (1.08-3.53) | 1.17 (0.90-1.52) |
| Prior diagnosed infection | None | Ref | Ref | Ref | Ref |
|  | 1st infection in 1st wave | 1.56 (0.50-4.92) | 2.63 (0.51-13.64) | 1.11 (0.14-78.87) | 2.12 (0.49-9.17) |
|  | 1st infection in 2nd wave | 0.32 (0.11-0.92) | 0.34 (0.10-1.09) | 0.68 (0.15-3.19) | 0.14 (0.03-0.67) |
| Vaccination status at time of diagnosis¶ | Unvaccinated | Ref | Ref | Ref | Ref |
|  | Partially vaccinated | 0.59 (0.45-0.77) | 0.79 (0.57-1.09) | 1.47 (0.32-6.66) | 0.57 (0.44-0.75) |
|  | Completely vaccinated | 0.15 (0.03-0.62) | 0.14 (0.02-1.20) | (omitted) | 0.22 (0.05-0.96) |

§ Adjusted for all variables in the model, as well as month of diagnosis, sub-district of residence and number of COVID-19 admissions in week of diagnosis.
\* Date of hospital admission within 21 days of COVID-19 date of diagnosis.
† The reference group here is the absence of that specific comorbidity.
¶ Fully vaccinated was defined as ≥28 days post-vaccination with Janssen/Johnson & Johnson or ≥14 days post second dose of Pfizer–BioNTech. Patients were deemed partially vaccinated from the day after their (first) vaccine dose until meeting criteria for complete vaccination.

